# The Trade-off Between Prioritization and Vaccination Speed Depends on Mitigation Measures

**DOI:** 10.1101/2021.02.24.21252352

**Authors:** Nikhil Agarwal, Andrew Komo, Chetan Patel, Parag Pathak, M. Utku Ünver

## Abstract

Calls for eliminating prioritization for SARS-CoV-2 vaccines are growing amid concerns that prioritization reduces vaccination speed. We use an SEIR model to study the effects of vaccination distribution on public health, comparing prioritization policy and speed under mitigation measures that are either eased during the vaccine rollout or sustained through the end of the pandemic period. NASEM’s recommended prioritization results in fewer deaths than no prioritization, but does not minimize total deaths. If mitigation measures are eased, abandoning NASEM will result in about 134,000 more deaths at 30 million vaccinations per month. Vaccination speed must be at least 53% higher under no prioritization to avoid increasing deaths. With sustained mitigation, discarding NASEM prioritization will result in 42,000 more deaths, requiring only a 26% increase in speed to hold deaths constant. Therefore, abandoning NASEM’s prioritization to increase vaccination speed without substantially increasing deaths may require sustained mitigation.

---

Vaccination can potentially alleviate the public health and economic crisis caused by SARS-CoV-2. The first vaccination in the US took place on December 14, 2020. As of February 19, 2021, the Pfizer-BioNTech and Moderna vaccines have been granted Emergency Use Authorization by the US Food and Drug Administration. About 59 million doses have been administered in two months, with 42 million individuals receiving at least one dose. It is expected to take 9 months to vaccinate 75% of the US population [1] and even longer for the world [2].

The scarcity of vaccines has motivated groups like the National Academies of Sciences, Engineering, and Medicine (NASEM) and the CDC’s Advisory Committee on Immunization Practices to propose allocation frameworks, taking into account age, social contacts, and other factors. US State, Tribal, Local, and Territorial entities (hereafter states) have used these recommendations as a basis for allocation.

Adherence to vaccine prioritization varies greatly amid concerns that implementing priorities is slowing down vaccination campaigns [3]. Some states have allowed lower priority patients to be vaccinated before those from earlier phases to increase speed and minimize waste [4], as vaccinations are seen as essential for the return to normalcy [5]. The fatigue and economic toll associated with sustaining social distancing, masking, and remote work creates a strong push towards faster vaccinations [6].

These forces have also motivated states to adopt mitigation policies that are responsive to the prevalence of infection [7, 8]. Many states are using a phased approach that relaxes or tightens restrictions based on health metrics. Under these policies, mitigation will depend on vaccine rollout. Ignoring this interaction could incorrectly predict disease dynamics.

The potential for increasing speed by abandoning prioritization motivates our investigation of how vaccine prioritization, vaccination speed, and mitigation measures affect cumulative mortality, herd immunity, years of life lost (YLL), and total incidence.

## Evaluating Disease Dynamics

We evaluated disease dynamics for SARS-CoV-2 under vaccine prioritization and mitigation scenarios using an age-stratified SEIR model, building on [9]. Age is a strong correlate of contact rates [10], susceptibility to infection [11, 12], and infection fatality rates [13, 14]. Moreover, recommended prioritization rules emphasize age after initial allocation phases [15]. We use eight age bins: 0-9, 10-19, …, 80+, and incorporate age-specific vaccine efficacy [16, 17] and vaccine hesitancy [18]. We forward simulate the path of the disease for one year starting from estimated initial conditions as of December 14, 2020. The initial number susceptible, infectious, and exposed in each age bin is calculated using death records from [19, 20]. To evaluate the effect of speed, we vary the number of individuals vaccinated per month. The assumptions and parameters of our model are detailed in the supplementary text.

We focus on two mitigation scenarios. Sustained Mitigation represents a partially-mitigated pandemic with a basic reproduction number *ℛ*_0_ of 1.5, which corresponds to an effective reproduction number *ℛ*_*t*_ of approximately 1.2 on December 14, 2020. This was the largest value for *ℛ*_*t*_ across all 50 states [21]. Assuming an unmitigated *ℛ*_0_ = 2.6 [22], it is equivalent to measures that scale contact rates by the factor *θ* = 0.577 until the end of the pandemic period. This factor *θ* measures the cumulative effect of all non-pharmaceutical interventions [23, 24]. (Figures S.6 – S.8 in the supplement provide results for Sustained Mitigation with *ℛ*_0_ = 1.3.)

Calibrated Mitigation represents time-varying mitigation measures that keep the total exposed and infectious population no higher than the initial level. Thus, the factor *θ* is a function of time that fixes *ℛ*_*t*_ at 1 until herd immunity is reached. This models a mitigation scenario that loosens measures as the force of infection decreases. Figure S.2 shows that *θ* starts at high mitigation (*θ ≈* 0.5) on Day 0 and moves towards no mitigation (*θ* = 1) at a rate that depends on vaccination policies. The approach of constraining *ℛ*_*t*_ at 1 follows [25].

We consider three vaccine prioritization policies: NASEM’s guidelines (NASEM), No Prioritization, and Optimal Prioritization. NASEM simulates the National Academies’ recommendation. After a “jumpstart” Phase 1A/B, NASEM’s Phase 2 allocates to K-12 staff and childcare workers, critical workers, individuals with comorbidities, older adults, and people in homeless shelters or prisons [15]. Our simulation of NASEM uses micro-level data with demographic and risk information for the US population [26]. No Prioritization corresponds to uniformly distributed vaccinations across the population. Optimal Prioritization minimizes cumulative number of deaths and depends on the mitigation scenario and vaccination speed.

It is optimal to vaccinate the elderly under both mitigation scenarios (Figure 1). In Optimal Prioritization, individuals over the age of 60, who have the highest IFRs and the lowest contact rates, are vaccinated before vaccinating Age 30-59. Vaccinated age groups overlap once a sufficient number of the older people have been vaccinated. Individuals below the age of 30 are not prioritized even though they have high contact rates.

**Figure 1:**
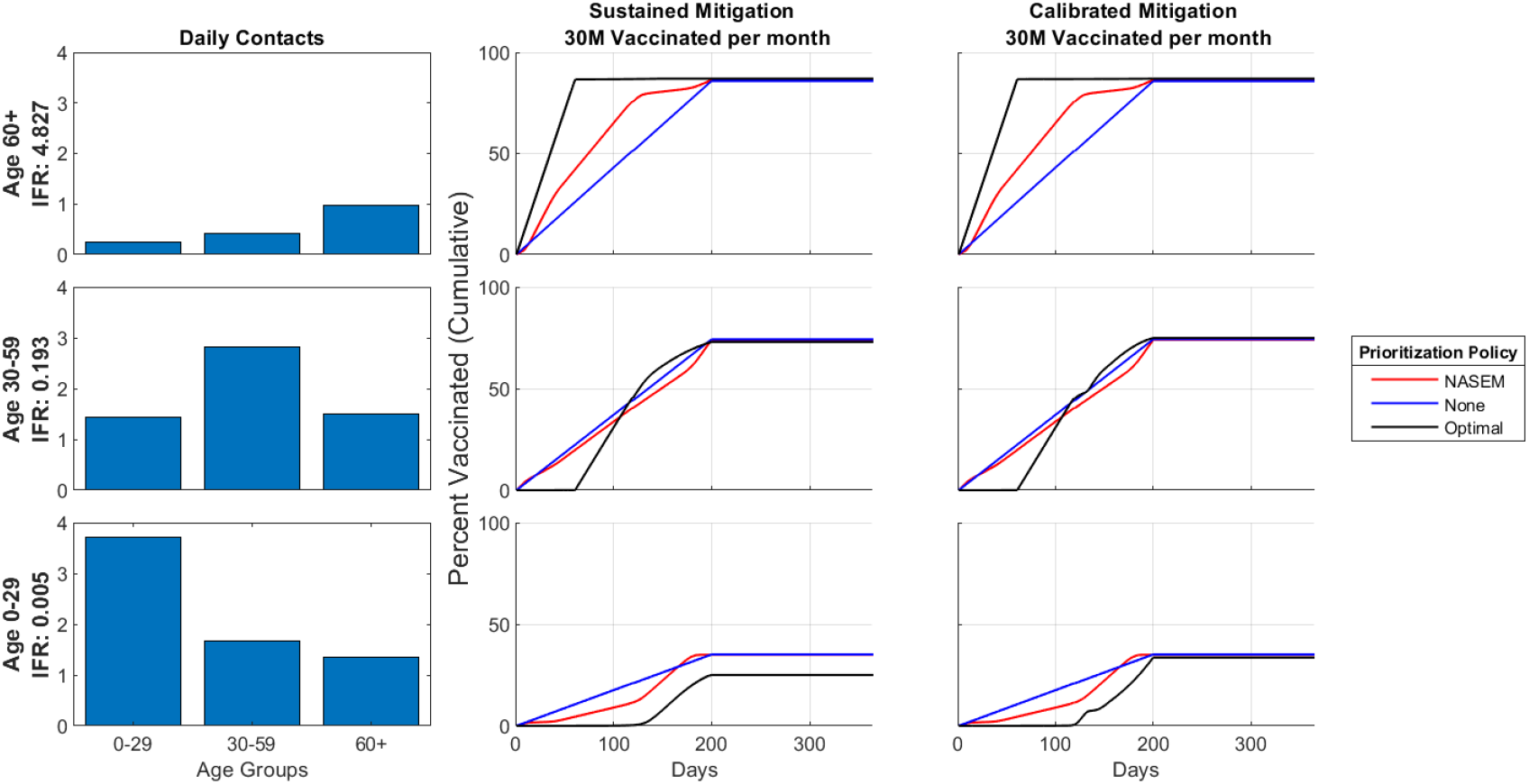
IFR, Contact Rates and Vaccination Policies by Age Group and Mitigation Scenario. Left panel shows IFR and contact rates aggregated to the age groups 0-29, 30-59, and 60+. Middle and right panels show vaccination policies for Sustained and Calibrated Mitigation with 30 million vaccinated per month.

NASEM’s recommendation also prioritizes the elderly, but not as much as the Optimal Prioritization. In NASEM, the vaccination of individuals less than 60 years begins alongside individuals over this age. However, NASEM vaccinates Age 60+ faster than No Prioritization, which vaccinates age groups in proportion to population.

These results focus on 30 million vaccinations per month (hereafter vpm) because that rate corresponds to the Federal government’s target to vaccinate every eligible individual by July 2021 [27], but are similar for 15 million vpm (Figures S.4, S.5).

## Impact of Prioritization and Mitigation on Disease Dynamics and Cumulative Deaths

Disease dynamics and cumulative deaths are sensitive to both prioritization policy and mitigation measures (Figure 2). Incidence first increases under Sustained Mitigation because initial *ℛ*_*t*_ *≈* 1.2, but rapidly declines within two months. Under Calibrated Mitigation, initial conditions result in momentum towards more infections for Age 60+ for a short period of time. However, the cumulative number of infections, deaths, and YLL are higher under Calibrated Mitigation than under Sustained Mitigation for each vaccine prioritization policy (Figure S.3).

**Figure 2:**
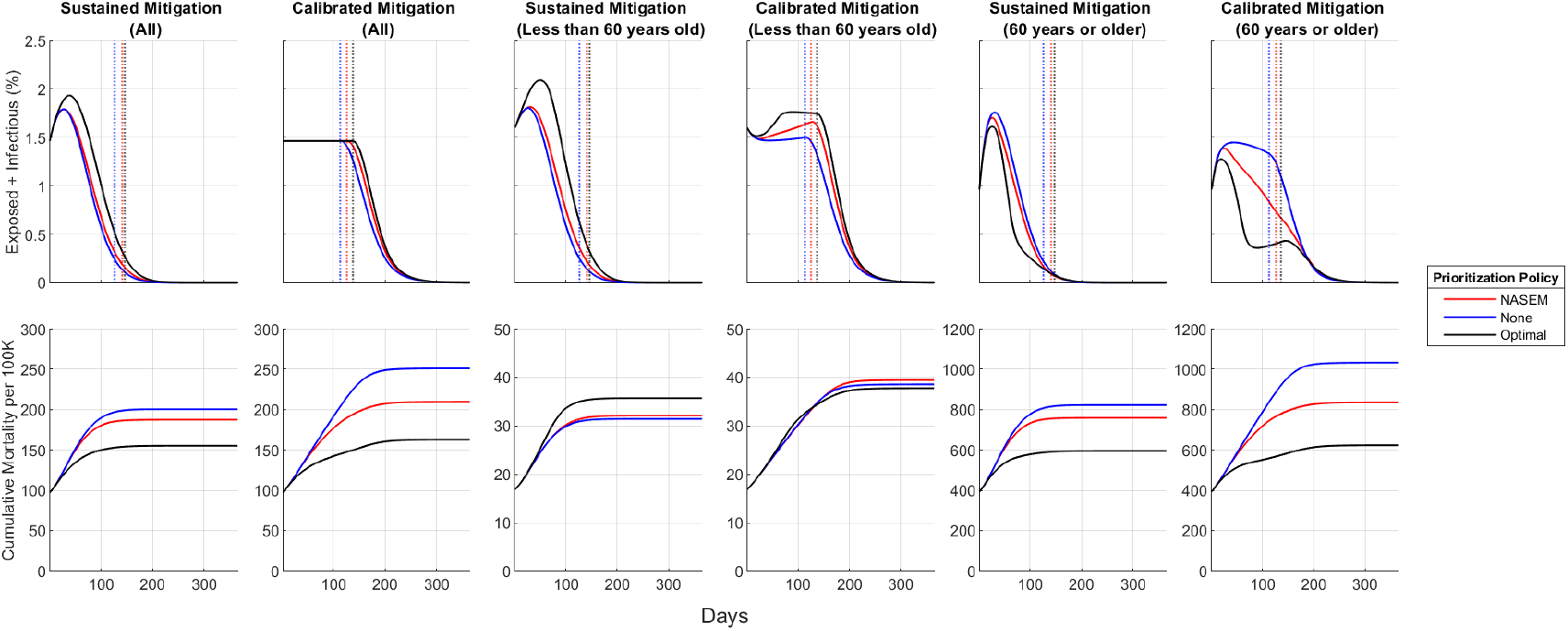
Active Cases and Cumulative Mortality over Time. Active cases (top panels) count both infectious and exposed individuals. Vertical dashed lines indicate time to herd immunity.

Although the total incidence is the highest under Optimal Prioritization, it saves the most lives (Figure 2). In both mitigation scenarios, Optimal Prioritization results in lower incidence for Age 60+ relative to the other prioritization policies, but higher incidence for Age 0-59. Cumulative deaths are between 104,000 and 151,000 greater under NASEM depending on the mitigation scenario. Relative to No Prioritization, NASEM similarly results in higher overall incidence, but lower incidence amongst the elderly and lower cumulative deaths. The comparisons are similar for YLL (Figures 2 and S.3).

These effects are driven by deaths for Age 60+ although this group comprises only 21.4% of the population. Under Optimal Prioritization, mortality for 60+ is between 623 and 695 per 100,000, but it is over 760 under other policies. In contrast, mortality for Age 0-59 is at most 40 per 100,000 under all scenarios.

## Effects of Vaccination Speed

We next evaluate the effect of vaccination speed on reduction of deaths and time to herd immunity relative to an unmitigated pandemic with no vaccinations (Figure 3). We also consider a No Mitigation scenario with *ℛ*_0_ = 2.6, which is calibrated to estimates in [22]. We consider speeds ranging from 15 million vpm to 40 million vpm. The former corresponds to 1 million doses per day of a two-dose vaccine, which is approximately equal to the rollout speed in the US at the end of the first month [28], to 40 million, which is higher than current Federal government targets [27].

**Figure 3:**
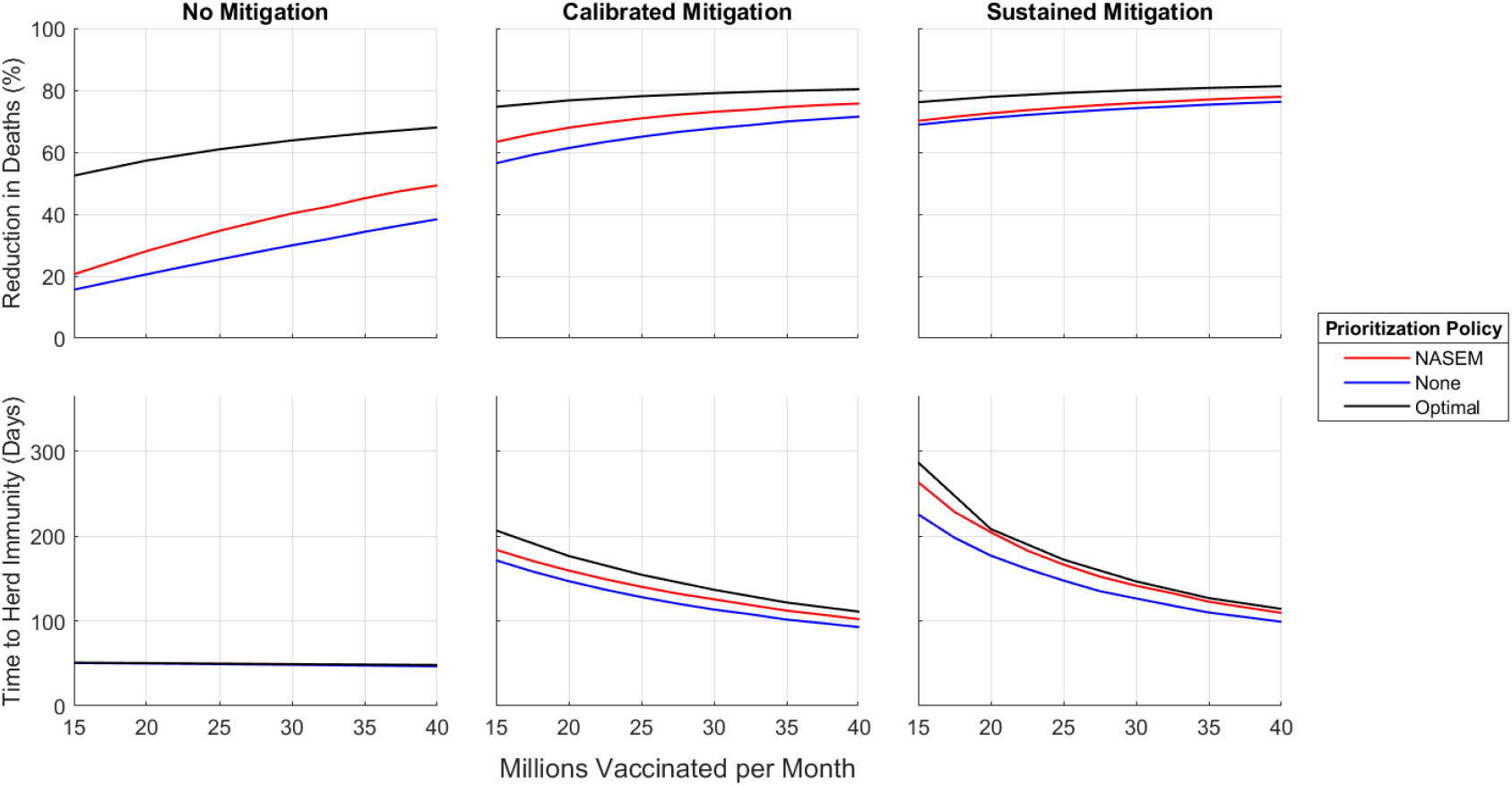
Reduction in Deaths and Time to Herd Immunity versus Vaccinations per month. The percentage reduction in deaths is compared to No Mitigation and no vaccines.

As expected, the benefits of increasing vaccination speed are large: it decreases both cumulative deaths and time to herd immunity under all scenarios considered. Increased mitigation also reduces cumulative deaths and delays herd immunity for a fixed vaccination speed and prioritization policy.

The reduction in deaths is larger under NASEM than under No Prioritization for all speeds and mitigation scenarios we considered. By construction, Optimal Prioritization outperforms both. These rankings are reversed for time to herd immunity. The differences between prioritization policies are more pronounced when mitigation measures are less stringent. For each level of speed, cumulative mortality across prioritization policies is most similar under Sustained Mitigation.

The benefits of marginal increases in vaccination speed also depend on the prioritization and mitigation scenarios. Increases in speed reduce cumulative deaths more if mitigation is lower: the effects on death are largest under No Mitigation, followed by Calibrated Mitigation. Given a mitigation scenario, cumulative deaths is more sensitive to vaccination speed under NASEM and No Prioritization than Optimal Prioritization. Across the range of scenarios considered, mitigation measures have an impact comparable to or larger than vaccination prioritization policies or vaccination speeds. Optimal Prioritization with 40 million vpm under No Mitigation results in no additional reduction in deaths than No Prioritization with 32.5 million vpm under Calibrated Mitigation or 15 million vpm under Sustained Mitigation. Similarly, the reduction in deaths under NASEM with 30 million vpm and Calibrated Mitigation is no higher than NASEM with 27.5 million vpm and Sustained Mitigation.

## The Trade-Off Between Prioritization and Speed

NASEM results in fewer cumulative deaths than No Prioritization for each speed and mitigation scenario considered (Figure 3). However, this ranking may be reversed if eliminating prioritization results in higher speed. Figure 4 analyzes this trade-off by reporting the increase in speed needed for No Prioritization to have the same cumulative deaths or YLL as NASEM. We present results varying speed and mitigation scenarios because Figure 3 shows that these two factors influence the effects of a marginal increase in speed.

**Figure 4:**
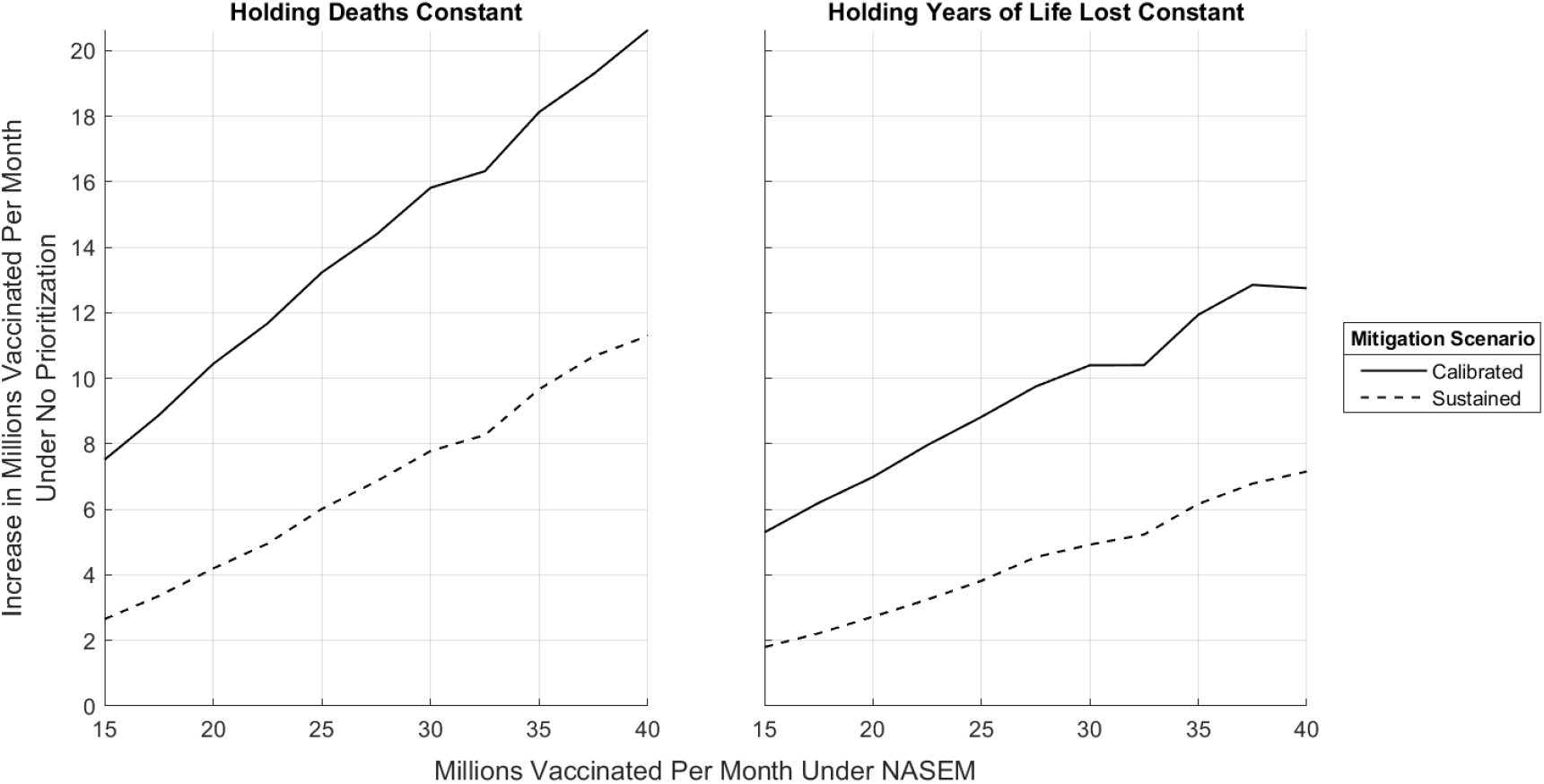
The Required Increase in Vaccinations per month under No Prioritization. Left panel keeps deaths the same as NASEM and plots the required increase relative to NASEM. Right panel keeps YLL the same as NASEM.

Compared to NASEM with Sustained Mitigation and 30 million vpm, No Prioritization would require an increase of at least 7.8 million vpm to achieve the same reduction in the number of deaths as NASEM. The required increase in speed is much larger under Calibrated Mitigation. At 30 million vpm, the minimum increase in vpm is 15.8 million, a 53% increase. These results are qualitatively similar if the comparison holds YLL constant, but smaller in magnitude. They are also robust to variations on the baseline assumptions which consider a vaccine with 80% efficacy (Figure S.10); set sustained mitigation so that *ℛ*_0_ = 1.3 (Figure S.8); or assume no vaccine hesitancy in the population (Figure S.9). Thus, abandoning NASEM’s prioritization policy in favor of No Prioritization while holding either deaths or YLL constant requires a larger increase in speed under Calibrated Mitigation.

## Discussion

Economic and political pressure may push states and individuals to relax mitigation measures as SARS-Cov-2 vaccination reduces infection and mortality. During initial vaccine rollouts, mask mandates and other mitigation efforts have been relaxed [29]. At the same time, current prioritization policies have been criticized as slowing vaccination speed [30]. This paper shows that relaxing restrictions during the rollout and abandoning vaccine prioritization could significantly increase mortality unless the gains in vaccination speed are dramatic.

We use an age-stratified SEIR model to benchmark vaccine prioritization policies under different mitigation scenarios. Across the scenarios, vaccinating elderly individuals who have the highest risk first minimizes cumulative deaths even though these individuals have the lowest contact rates (see also [9]). NASEM guidelines include prioritization for the elderly, but less so than Optimal Prioritization in part due to ethical considerations for healthcare and essential workers [15]. In fact, Optimal Prioritization illustrates that finer age-based prioritization could be valuable even after initial vaccination phases. No Prioritization does not target high-risk individuals, increasing deaths.

Although NASEM reduces deaths relative to No Prioritization, the differences between these policies are significantly smaller if the spread of infections is suppressed by continuing mitigation measures during vaccine distribution. A more realistic mitigation approach that gradually relaxes restrictions during the vaccine distribution results in a larger difference in deaths between NASEM and No Prioritization. Nonetheless, abandoning NASEM may not increase deaths if No Prioritization results in higher vaccination speed. The required gain in speed to keep deaths from increasing is smaller if mitigation measures can be sustained. Thus, combining vaccine prioritization policy with mitigation scenarios in SEIR models is essential for understanding disease dynamics and the health effects of vaccination.

The focal outcomes we study are cumulative deaths and time to herd immunity. The ranking between the prioritization strategies and the qualitative trade-off between speed and prioritization are similar with YLL. However, infection rates are higher and herd immunity is delayed under prioritization policies that reduce deaths. The strain on healthcare capacity or economic costs may weigh in favor of prioritization policies that instead reduce infections. It is possible to adapt our framework to compare the effects of vaccine prioritization and specific mitigation measures on healthcare utilization [31] or economic impacts [24].

Several states have adapted the NASEM benchmark in devising their own guidelines. Similarly, Calibrated Mitigation is a stylized depiction of the phased approach to restrictions being used in several states. Enriching our framework to more accurately represent vaccine prioritization policies and mitigation scenarios requires more detailed information on these policies. With this information, our model can forecast the interplay between prioritization, mitigation, and speed for particular states.

Our NASEM simulation uses micro data from the US population. Detailed estimates of epidemiological factors that vary by demographics and occupation could be used to further stratify our SEIR model. For instance, contact rates and adherence to mitigation may vary by these characteristics. The model could use such information to more precisely evaluate the effect of prioritizing healthcare workers and first responders in Phase 1A, which represent 5% of the U.S. population. Many guidelines recommend prioritizing these groups partly because of their specific risks and contact rates. Nonetheless, our age-stratified model captures the vast majority of vaccinations occurring after the initial phase.

Our study has several important limitations. Predicting the course of the disease presents challenges for modeling vaccine prioritization policy because of high uncertainty. For example, it is unknown whether recovered individuals have enduring immunity [32]. There are also new strains of SARS-Cov-2, with differing levels of transmissibility and virulence [33, 34], and vaccine efficacy [35, 36, 37, 38]. How these new strains affect the consequences of vaccine prioritization is unclear. Higher virulence increases the importance of age-based prioritization, vaccination speed, and mitigation. While our model abstracts away from these new strains, understanding the trade-off between prioritization, speed, and mitigation will be necessary for designing effective vaccination and mitigation policies as the disease evolves.

## Data Availability

Available at the below link along with replication code.

https://github.com/patelchetana/vaccine-speed-vs-prioritization

## Supplementary Text

### Materials and Methods

#### The Susceptible Exposed Infectious Recovered (SEIR) Model with Vaccinations and Deaths

We use a continuous-time ordinary differential equation (ODE) model with age-stratified compartments (*S*_*i*_, *E*_*i*_, *I*_*i*_, *R*_*i*_, *D*_*i*_, *V*_*i*_) indexed by *i* in the age-groups 0-9, 10-19, …, 70-79, 80+. The model builds on [9] by incorporating the initial state of the pandemic as of December 14, 2020; the vaccine prioritization strategy recommended by the National Academies of Sciences, Engineering and Medicine (NASEM) [15]; and a calibrated community mitigation scenario.

Initially susceptible individuals (state *S*) may transition to the exposed state (*E*) upon contact with infectious (*I*) individuals. Exposed individuals transition to the infectious state, after an expected duration of *d*_*E*_. Infectious individuals, after an expected duration of *d*_*I*_, transition to either the recovered (*R*) or the dead (*D*) state with probability given by age-specific IFR from [13]. We assume that recovered individuals are immune from infection. The expected durations *d*_*E*_ and *d*_*I*_ are age-invariant and calibrated based on [39, 40]. The number of individuals in group *i*, denoted *N*_*i*_, is calculated by scaling the 2014-2018 5-year American Community Survey (ACS) Public Use Micro Data [41] to its full population representation using the included person weights.

We allow susceptible, exposed, and recovered individuals to be vaccinated because differentiating between these groups has not been recommended in the US [15]. However, vaccinations move only susceptible individuals to the (*V*) state. Vaccines do not alter the course of individuals already exposed when immunized. We assume that vaccines are transmission blocking if they protect an individual; immunity, if acquired through vaccination, is immediate once the vaccination course has been completed; and no individual can be vaccinated more than once. For two-dose vaccines like the Pfizer-BioNTech and Moderna vaccines, a course is completed only after both doses have been administered.

Vaccine efficacy is introduced by adding compartments *S*_*V*_, *E*_*V*_, *I*_*V*_ and *R*_*V*_. Susceptible individuals transition to the state *S*_*V*_ upon vaccination with probability *ν*_*e,i*_, which is calibrated to age-specific efficacy measured in [16, 17]. Individuals in *S*_*V*_ transition to *E*_*V*_ and *I*_*V*_ according to the same laws as unvaccinated individuals. However, consistent with evidence from clinical trials, all vaccinated individuals are fully protected from death [16, 17].

We assume that community mitigation scales contact rates proportionally across all groups. Thus, the force of infection for group *i* at time *t* is given by

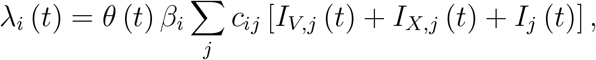

where *θ* (*t*) controls the cumulative impact of all community mitigation measures at time *t, β*_*i*_ is the transmission probability following a contact with an infectious individual, *c*_*ij*_ is the number of daily contacts for individuals in group *j* that an individual in group *i* contacts, and *I*_*j*_ (*t*), *I*_*X,j*_ (*t*) and *I*_*V,j*_ (*t*) are the proportions of individuals in group *j* that are infectious and respectively (i) unvaccinated, (ii) vaccine hesitant, and (iii) vaccinated but unprotected from infection. Thus, the sum of these terms is the probability that an individual in group *j* is infectious.

We project age-specific contacts rates using data from [10] to the US population to calculate *c*_*ij*_. We use a US-specific contact matrix identifying interactions in all available location types (rural and urban) and settings (home, work, school, and other). We follow the approach of [9] to collapse 5-year age groups into the 10-year groups in our model. We depart from [9] by using population weights from the ACS. We again follow [9] to extend contact matrices to include individuals aged 80+ by copying the contact rates for individuals aged 70-79 along the diagonal and then adjusting 80+ contact rates to account for long-term care facility interactions. Specifically, contacts between 80+ individuals and 0-60 year old individuals are scaled down by 10% with the amount decreased then evenly redistributed to interactions with 70-79 and 80+ year olds.

The basic reproduction number is *ℛ*_0_, which is the first eigenvalue of the next-generation matrix without mitigation *D*_*β*_*CD*_*I*_, where *D*_*β*_ is the diagonal matrix with elements *β*_*i*_, *D*_*I*_ is the diagonal matrix with elements *d*_*I,i*_, and *C* is a matrix with *i − j* element *c*_*ij*_. We assume that *β*_*i*_ is proportional to age-specific susceptibility estimated in [12] (see Table S.3). The vector *β* is scaled to calibrate *ℛ*_0_ to 2.6, which is the midpoint of values estimated in [22].

Figure S.1 presents a schematic description of the model discussed above. Table S.3 presents the calibrated parameters and Table S.1 presents the contact matrix.

#### Initial Conditions

Initial conditions for the model as of December 14, 2020 are calculated using data on the age-distribution of reported deaths sourced from the Centers for Disease Control and Prevention (CDC) [19] and smoothed state-level death counts over time compiled by the New York Times [20], IFR rates in [13], and *d*_*E*_ and *d*_*I*_. The number initially vaccinated is zero. Following [42], we combine estimates from [40] of incubation period and time from symptom onset to death to construct an uncertainty interval of lag of deaths to exposure of 18 to 24 days. We assume that lags are discretely uniformly distributed following [43]. This gives estimates of the true (not reported) number of new exposures per day. The number of initially susceptible individuals is obtained by subtracting the cumulative number of individuals ever exposed until December 14, 2020 from the number of individuals in the corresponding group, *N*_*i*_. The number of individuals initially exposed is the number of estimated new exposures over the course of the *d*_*E*_ days prior to December 14, 2020. The number of infectious individuals on December 14, 2020 is the number of estimated newly exposed individuals over the course of the *d*_*I*_ days preceding *d*_*E*_ days prior to December 14, 2020. The number of initial recovered or dead individuals is the cumulative number of estimated exposures through the date *d*_*I*_ + *d*_*E*_ + 1 days prior to December 14, 2020. Throughout our calculations of initial conditions, we assume that no individual has been infected twice.

Table S.2 presents our initial conditions. Table S.4 compares our imputed number of total true infections against various sources in the literature. While several studies have found that Covid-19 reported case counts underestimate actual infections, there is limited consensus in the literature on the true disease incidence of Covid-19. Our estimates fall in the middle of the tabulated total number of true infections. Relative to [44] and [45], our estimates of total true infections are moderately higher. Our estimates are in line with those of [46] and underestimate relative to [47]. Table S.5 compares our age-stratified imputation against the CDC estimates [48]. The CDC estimates are through the end of December while ours are through December 14, 2020. We are therefore unable to directly compare the two true infection estimates. However, the age-distribution of cumulative infections as reported by the CDC closely matches the age-distribution of our total number of infections.

#### Sustained and Calibrated Mitigation Scenarios

We simulate two types of mitigation scenarios. Sustained Mitigation sets *θ* (*t*) = *θ*_0_ for all *t*, with *θ*_0_ = 1.5*/*2.6. This value yields an effective reproduction number *ℛ*_*t*_ on the initial date of approximately 1.2 since 79.4% of the population is initially susceptible on December 14, 2020 according to our estimates (see Table S.4). The model is equivalent to setting *ℛ*_0_ = 1.5 and evaluating it under no mitigation, that is, *θ*_0_ = 1.

The Calibrated Mitigation scenario sets *θ* (*t*) *∈* [0, 1] so that the total exposed plus infectious remains constant until herd immunity is attained. The value of *θ* (*t*) is the solution to the problem

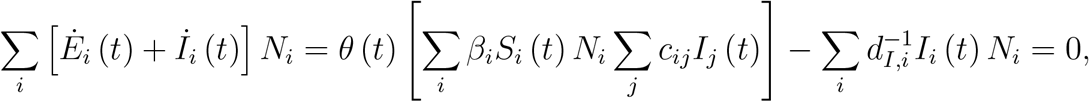

where we have combined the compartments *E*_*V*_ and *E*_*X*_ into *E*, and *I*_*V*_ and *I*_*X*_ into *I* for simplicity of notation. Thus, this sets *ℛ*_*t*_ = 1 until herd immunity is attained since the expected number of new exposures caused by a newly infectious individual at time *t* is equal to 1.

#### Time to Herd Immunity

Time to herd immunity is defined as the earliest date on which the total infectious plus exposed would drop if mitigation measures were lifted:

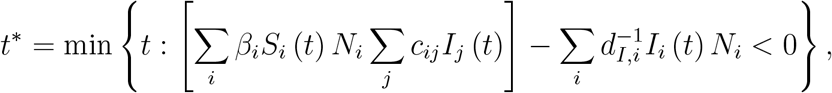

where we have combined the compartments *E*_*V*_ and *E*_*X*_ into *E*, and *I*_*V*_ and *I*_*X*_ into *I* for simplicity of notation.

#### Incorporating Vaccination Hesitancy

We incorporate vaccine hesitancy by introducing compartments *S*_*X*_, *E*_*X*_, *I*_*X*_ and *ℛ*_*X*_. Individuals are placed in *S*_*X*_ if they are hesitant and may only transition to *E*_*X*_, *I*_*X*_, *ℛ*_*X*_ and *D*. The proportion of individuals placed in *S*_*X*_ is age-specific and calibrated using a January 2021 survey administered by the U.S. Census Bureau [18]. Vaccine uptake rates, equal to one minus hesitancy, are defined as the age-stratified proportion of individuals responding to the Census survey as: (i) already having received at least one vaccine dose, (ii) indicating the individual will probably get the vaccine, or (iii) indicating the individual will definitely get the vaccine. The Census age stratification does not match the model group definitions. Thus, we translate their estimates to the model group structure using population weights from the ACS data. Further, consistent with the limitations on the authorization of currently available vaccines, we assume that individuals below the age of 16 cannot be vaccinated [49, 50]. This is incorporated into our model by assuming that their uptake is 0% and thus they are fully hesitant.

#### Simulating the NASEM Vaccination Policy

The NASEM guidelines define a phased allocation meant to inform vaccine prioritization policy at the federal, state, and local level. They outline five phases (1A, 1B, 2, 3, 4) based on a combination of demographics, individual health risk, and individual occupation. We label each individual in the ACS data to the highest priority NASEM phase for which they qualify and simulate an iterative federal to state to individual allocation of vaccines. Federal allocation of incrementally available units to states is by state population share. State allocation of received units to individuals is in NASEM phase order where within-phase allocation proceeds by random lottery. Our phase-labeling procedure follows that of [26] and is outlined below.

Phase 1A includes healthcare workers in high-risk settings and first responders. We identify these health care workers through industry codes from the ACS data. NASEM also includes death care professionals, pharmacists, public health workers, and dentists alongside frontline health care workers. These individuals, along with the first responders, are identified using occupation codes. We use the analysis from [51] to only include health care workers in high-risk settings.

Phase 1B includes individuals of all ages with health conditions that put them at significantly higher risk. We use the CDC 2018 Behavioral Risk Factor Surveillance System (BRFSS) dataset to assess risk and merge it with the ACS [52]. For each group defined by age, sex, and race in the BRFSS data, we compute the proportion of individuals with at least two risky health conditions. The BRFSS does not include data on individuals younger than 18, therefore risk probabilities of the youngest age-bin with available data, 18-24, are extrapolated to individuals younger than 18. For each observation in the ACS data, we assign individuals to high risk based on the risk probabilities computed from the BRFSS conditional on demographics. Phase 1B also includes older adults living in congregate or overcrowded settings. We identify these individuals in the ACS data by including individuals who are at least 65 years old and live in multigenerational housing or institutional group quarters.

Phase 2 guidelines include several groups. First, K-12 teachers, school staff, and child care workers are identified using occupation codes. Second, critical workers working in settings of high exposure-risk are identified using LMI Institute coding of the Department of Homeland Security (DHS) definition of Essential Critical Infrastructure Workers to identify critical workers [53]. High exposure-risk is computed using survey results from the Bureau of Labor Statistics (BLS) O*NET on disease exposure in the workplace [54]. Third, individuals of all ages with at least one risky health condition are identified using the same methods as in Phase 1B. Fourth, people living in homeless shelters or group homes as well as staff who work in such settings are identified by including people living in non-institutional group quarters according to the ACS data. As non-institutional group quarters also include military barracks and college dormitories, we remove any active military members and undergraduate college students from consideration in non-institutional group quarters [55]. Staff in such settings are identified by occupation code. Fourth, prisoners and prison staff are identified as individuals in institutional group quarters and occupation codes are used to identify prison staff. Finally, we include all individuals at least 65 years old.

Phase 3 includes young adults, children, and critical workers not included in Phase 1 or 2. Per NASEM guidelines, we include young adults as anyone at least 18 years old and at most 30 years old. We include children by adding everyone less than 18 years old. Using the same critical worker definition from Phase 2, we include all critical workers who have not yet been assigned a phase.

Finally, we assign anyone remaining unassigned to Phase 4.

The NASEM guidelines contain approximate estimates of phase sizes. Our phase labeling procedure yields phase sizes that closely match their estimates. Specifically, they specify Phase 1A as 5%, Phase 1B as 10%, Phase 2 as 30-35%, Phase 3 as 40-45%, and Phase 4 as 5-15% of the US population. Our labeling yields Phase 1A as 5%, Phase 1B as 11%, Phase 2 as 37%, Phase 3 as 39%, and Phase 4 as 8% of the US population.

#### Solving the Model

We simulate the model by solving the ODE using a daily discrete-time approximation for 365 days. The pandemic period ends by that time in our simulations. The optimal vaccination policy *{V*_*i*_ (*t*)*}*_*i*_ is a function of age-group and time. We solved for this policy using Artleys’ KNITRO optimization software. The problem is constrained so that the total daily flow of vaccinated individuals across all groups 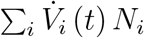 cannot exceed the total number of vaccination courses completed each day. The optimizer is initialized at an allocation proportional to the initial population share in each age group.

**Figure S.1:**
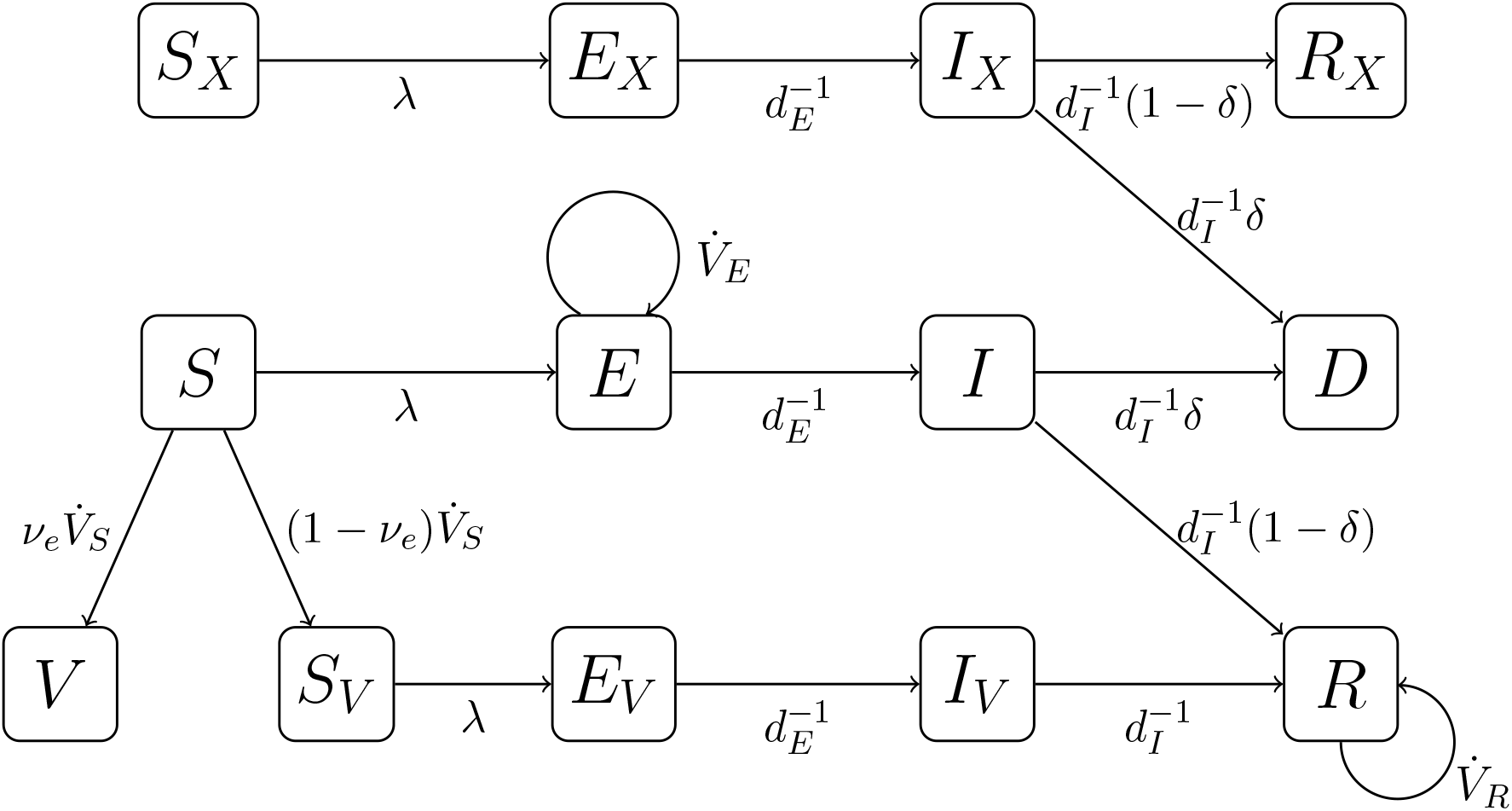
Schematic Depiction of the SEIR Model. *S*_*X*_, *S*_*V*_, and *S* are susceptible; *E*_*X*_, *E*, and *E*_*V*_ are exposed; *I*_*X*_, *I*, and *I*_*V*_ are infectious; *ℛ*_*X*_ and *ℛ* are recovered; *D* is dead; *V* is vaccinated and protected. The subscripts *X* and *V* denote vaccine hesitant and vaccinated but unprotected respectively. 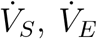 and 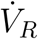 are the rates at which susceptible, exposed and recovered individuals are vaccinated. We ensure that no individual is vaccinated twice by moving individuals from *E* and *ℛ* to separate compartments at rates 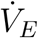 and 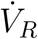 respectively. *ν*_*e*_ denotes vaccine efficacy; *λ* denotes the force of infection; *d*_*E*_ and *d*_*I*_ are the expected durations in the exposed and infectious states respectively; *δ* is the IFR.

**Table S.1:**
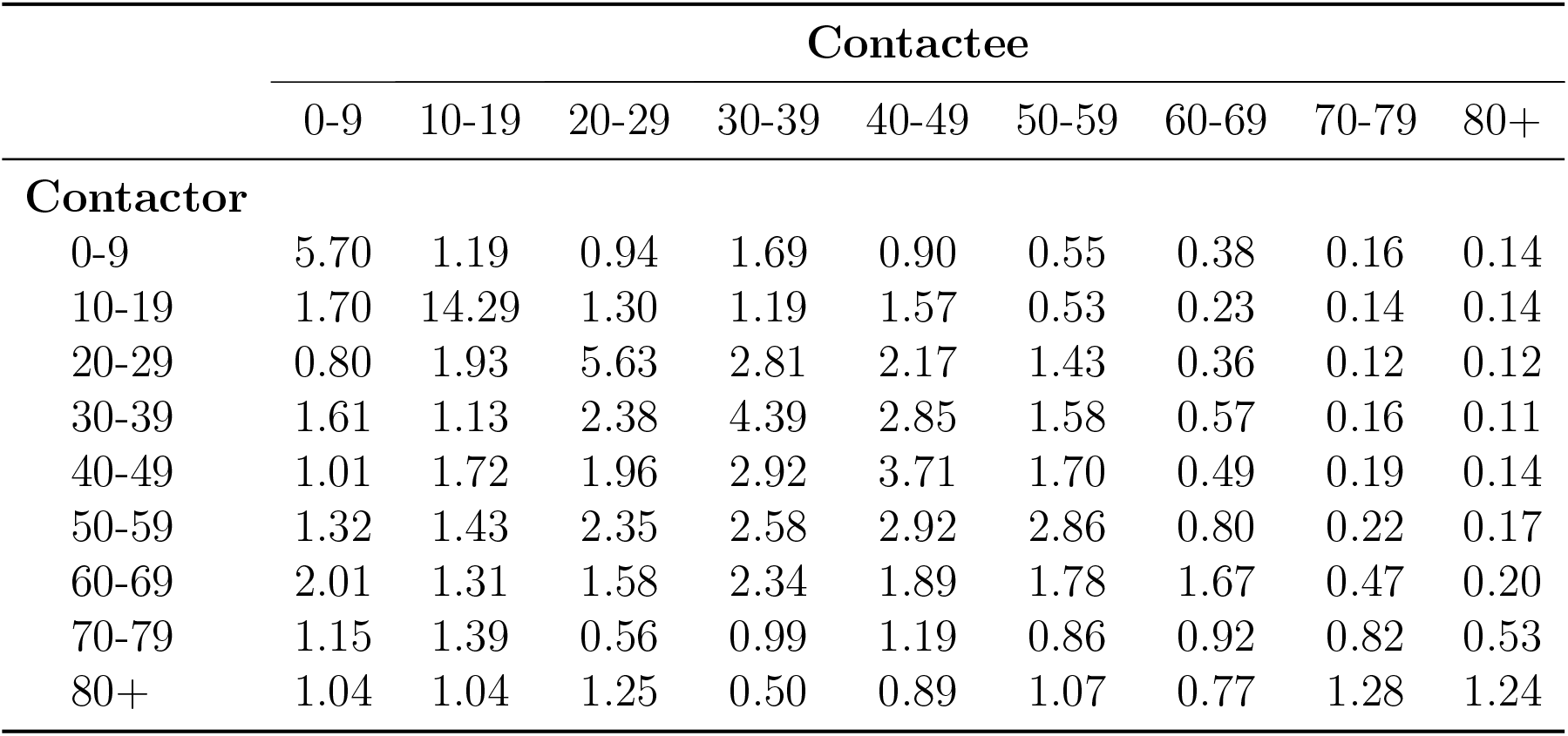
Mean daily contacts by age group. Mean daily contacts by age group in the United States in all location types and settings from [10]. Conversion from 5-year to 10-year age intervals and extrapolation to include 80+ individuals following [9].

**Table S.2:** Initial Conditions by Age Group as of December 14, 2020. Age-stratified share of population in each model compartment at start of simulation. *S*(0) is the share susceptible in the *S, S*_*X*_ and *S*_*V*_ groups combined; *E*(0) is the share exposed in the *E, E*_*X*_ and *E*_*V*_ groups combined; *I*(0), share infectious in the *I, I*_*X*_ and *I*_*V*_ groups combined; *ℛ*(0) is share recovered in the *ℛ* and the *ℛ*_*X*_ groups; *D*(0) is the share dead.

| | $S(0)$ | $E(0)$ | $I(0)$ | $R(0)$ | $D(0)$ |
| --- | --- | --- | --- | --- | --- |
| **Age group** |  |  |  |  |  |
| 0-9 | 0.7705 | 0.0051 | 0.0105 | 0.2139 | <0.0001 |
| 10-19 | 0.7709 | 0.0044 | 0.0092 | 0.2156 | <0.0001 |
| 20-29 | 0.7094 | 0.0060 | 0.0122 | 0.2724 | <0.0001 |
| 30-39 | 0.7368 | 0.0058 | 0.0123 | 0.2451 | 0.0001 |
| 40-49 | 0.7834 | 0.0053 | 0.0115 | 0.1996 | 0.0002 |
| 50-59 | 0.8346 | 0.0044 | 0.0094 | 0.1510 | 0.0006 |
| 60-69 | 0.8763 | 0.0035 | 0.0075 | 0.1113 | 0.0015 |
| 70-79 | 0.9087 | 0.0027 | 0.0058 | 0.0792 | 0.0036 |
| 80+ | 0.9199 | 0.0024 | 0.0052 | 0.0621 | 0.0104 |

**Table S.3:** Calibrated model parameters. Summary of age-stratified parameters used in model and simulation. *d*_*E*_, length of exposed period; *d*_*I*_, length of infectious period; *β*_*i*_ susceptibility to infection for age group *i*; *IFℛ*_*i*_, infection fatality rate for age group *i*; *N*_*i*_, number of people in age group *i*; *ν*_*u,i*_, vaccine uptake (one minus vaccine hesitancy) for age group *i*; *c*_*ij*_, contact rate between age group *i* and *j*; *ν*_*e,i*_, vaccine efficacy among age group *i*; *Y LL*_*i*_, years of life lost upon each death in age group *i*. Vaccine efficacy represents an unweighted combination of Moderna and Pfizer age-stratified efficacy. Efficacy is extrapolated to children using reported efficacy among youngest subgroup. Vaccine uptake is assumed to be 0% for individuals less than 16 years old. ACS population data is used to align age groups from original efficacy and uptake sources to the groups in the model. Years of life lost are summarized at the age group level using population weights from the ACS.

|  | Age group |  |  |  |  |  |  |  |  | Source |
| --- | --- | --- | --- | --- | --- | --- | --- | --- | --- | --- |
|  | 0-9 | 10-19 | 20-29 | 30-39 | 40-49 | 50-59 | 60-69 | 70-79 | 80+ |  |
| $d_E$ | 4 days (age invariant) | | | | | | | | | [39, 40] |
| $d_I$ | 9 days (age invariant) | | | | | | | | | [39] |
| $\beta_i$ | 0.40 | 0.38 | 0.79 | 0.86 | 0.80 | 0.82 | 0.88 | 0.74 | 0.74 | [12] |
| $IFR_i$ | 0.001 | 0.003 | 0.011 | 0.037 | 0.123 | 0.413 | 1.380 | 4.620 | 15.460 | [13] |
| $N_i$ | 40,065,319 | 42,116,480 | 45,002,935 | 42,518,642 | 40,909,501 | 43,310,372 | 36,150,682 | 20,706,539 | 12,122,561 | [41] |
| $\nu_{u,i}$ | 0.000 | 0.298 | 0.724 | 0.711 | 0.750 | 0.789 | 0.858 | 0.893 | 0.893 | [18] |
| $c_{ij}$ | See Table S.1 | | | | | | | | | [10, 9] |
| $\nu_{e,i}$ | 0.956 | 0.956 | 0.956 | 0.956 | 0.956 | 0.951 | 0.922 | 0.911 | 0.932 | [17, 16] |
| $YLL_i$ | 74.5 | 64.7 | 55.1 | 45.9 | 36.6 | 27.8 | 20.0 | 13.0 | 6.5 | [56] |

**Table S.4:**
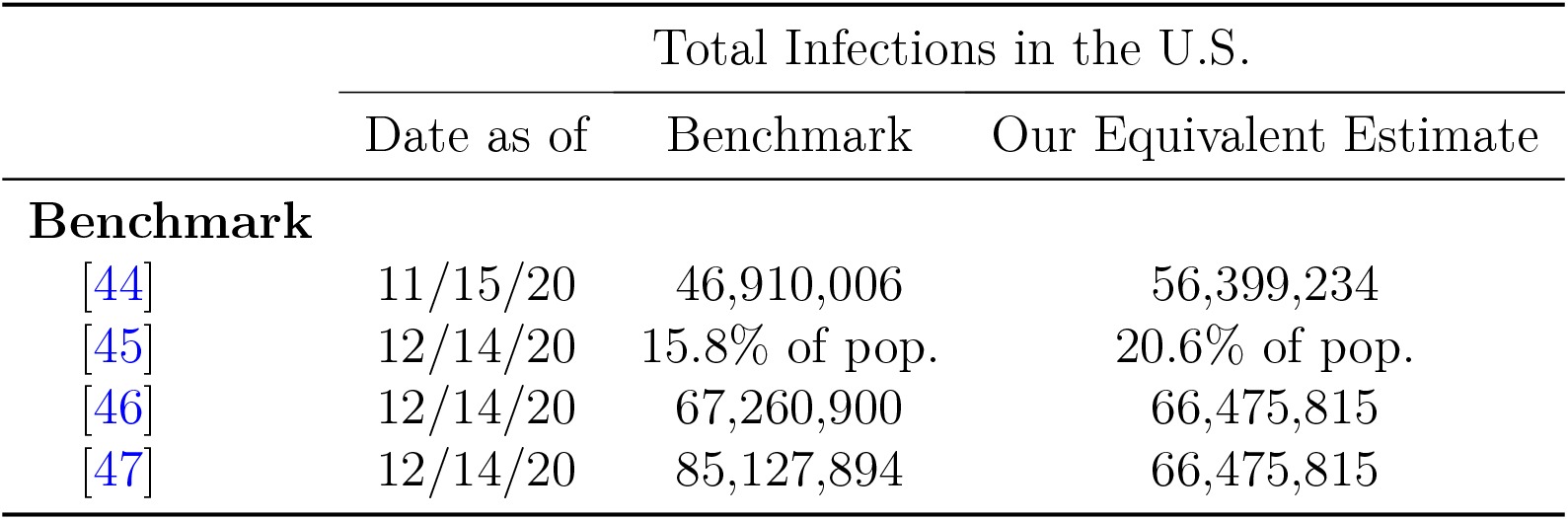
Imputed cumulative infections by source. [46] estimates are as reported on February 16, 2021. [47] estimates are as reported on December 20, 2020 for their maintain status quo scenario. Our imputation of the percent of US population that has been infected is computed using population counts from the ACS.

|  | Total Infections in the U.S. |  |  |
| --- | --- | --- | --- |
|  | Date as of | Benchmark | Our Equivalent Estimate |
| **Benchmark** |  |  |  |
| [44] | 11/15/20 | 46,910,006 | 56,399,234 |
| [45] | 12/14/20 | 15.8% of pop. | 20.6% of pop. |
| [46] | 12/14/20 | 67,260,900 | 66,475,815 |
| [47] | 12/14/20 | 85,127,894 | 66,475,815 |

**Figure S.2:**
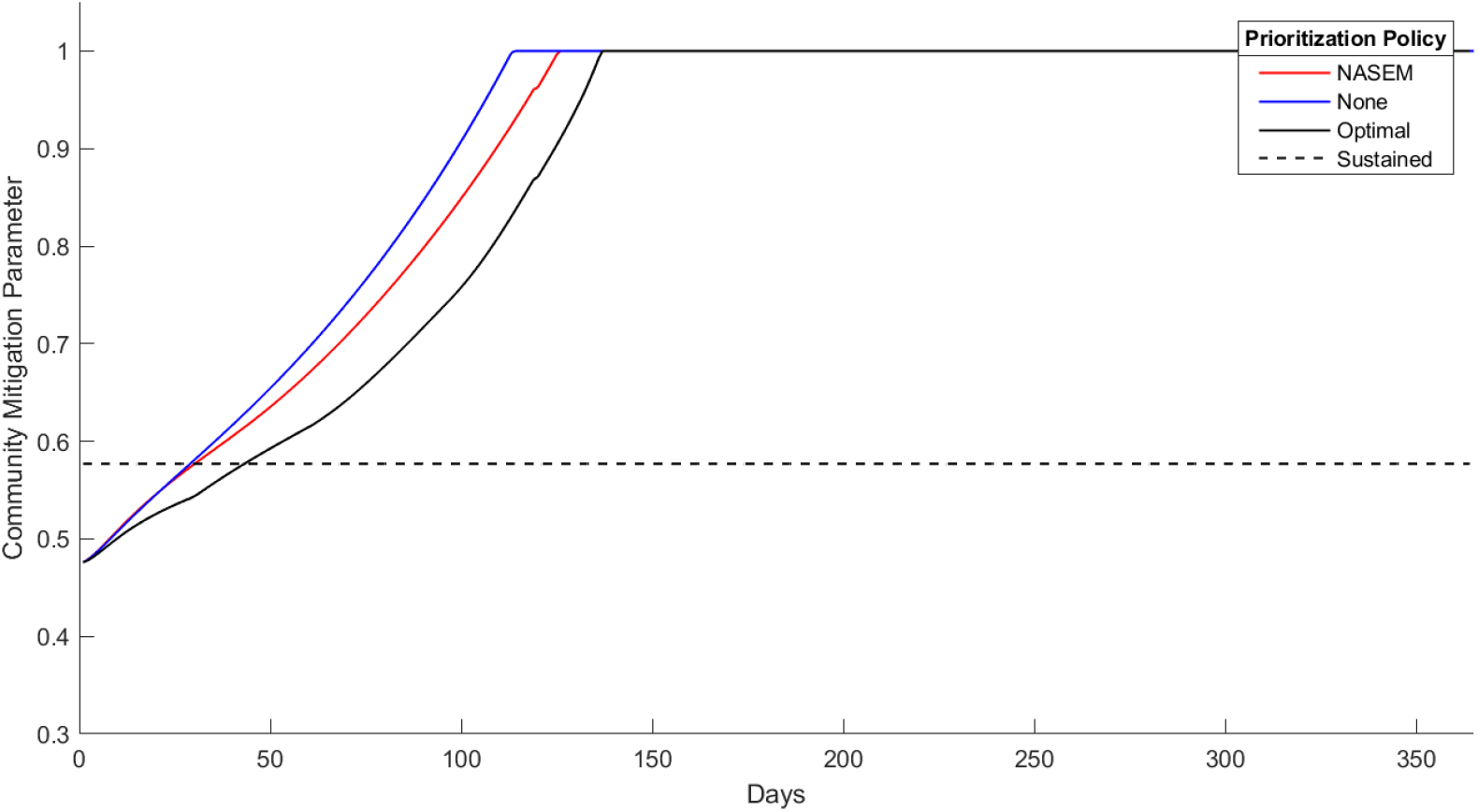
Mitigation Measures Over Time. The parameter *θ* over time under Calibrated Mitigation and Sustained Mitigation (with *ℛ*_0_ = 1.5). The mitigation parameter scales the force of infection proportionally for each group. Lower values indicate more stringent measures.

**Figure S.3:**
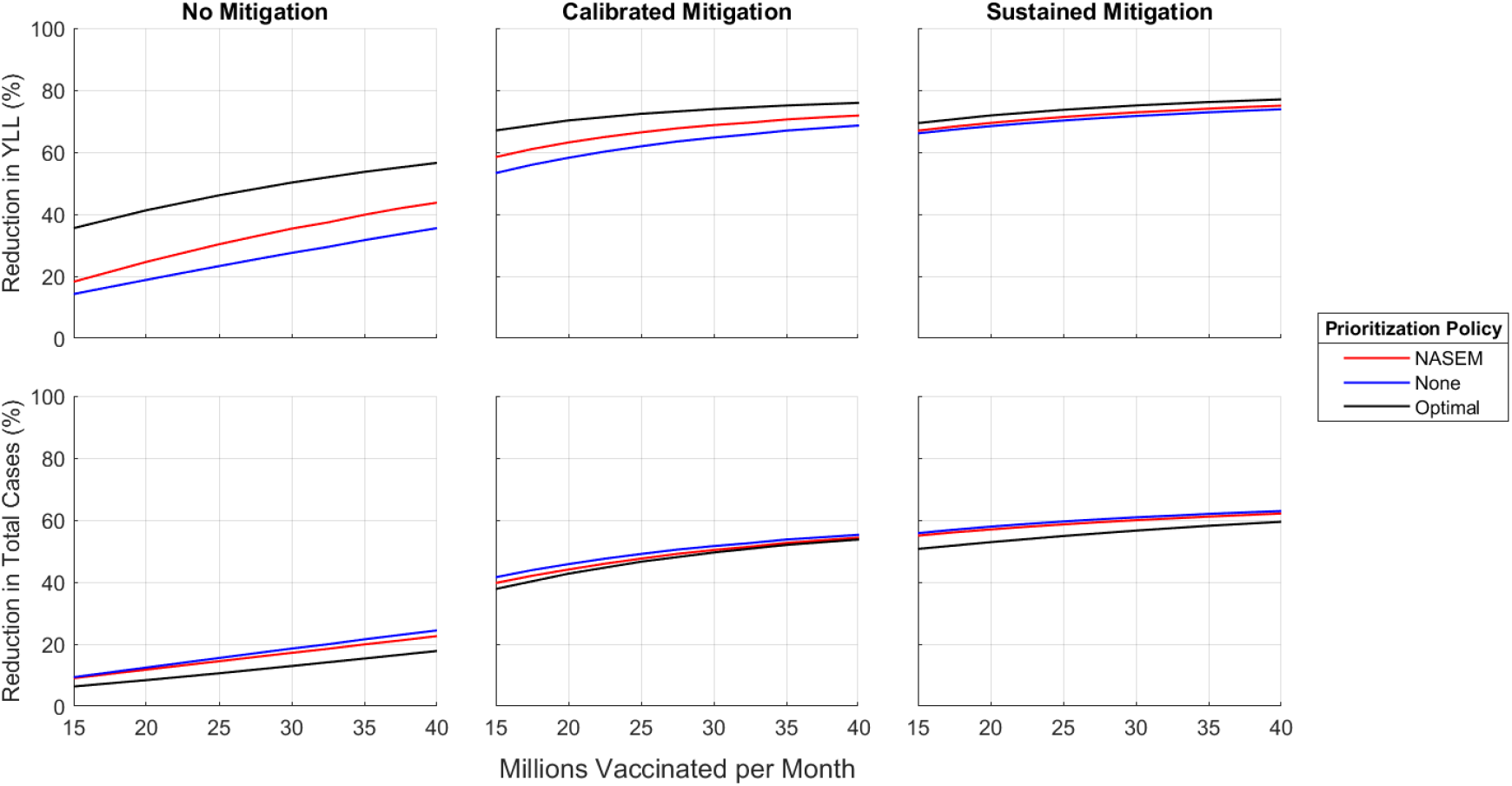
Reduction in YLL and Total Cases versus Vaccinations per Month. The percentage reductions compared to No Mitigation and no vaccines.

**Figure S.4:**
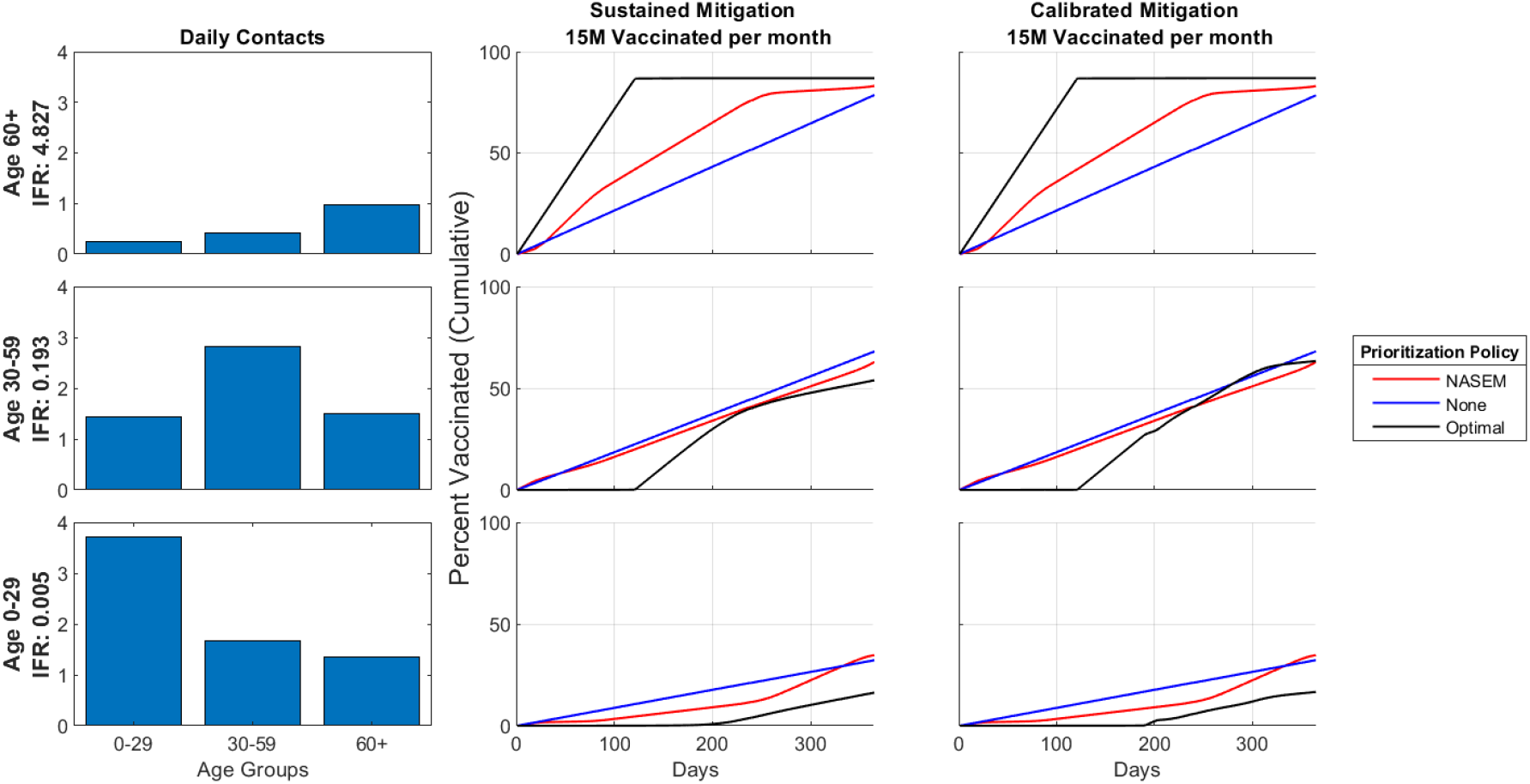
IFR, Contact Rates and Vaccination Policies by Age Group and Mitigation Scenario for 15 million vaccinated per month. Analogous to Figure 1.

**Figure S.5:**
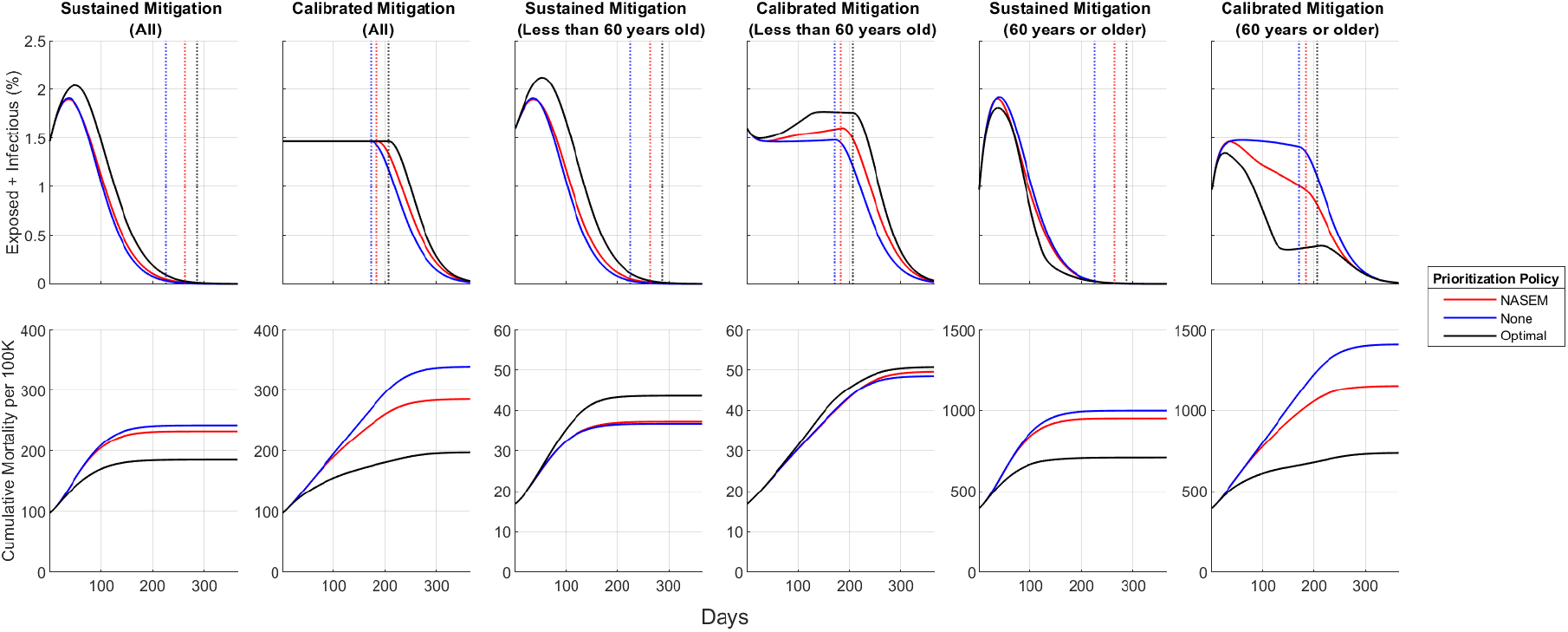
Active Cases and Cumulative Mortality over Time for 15 million vaccinated per month. Analogous to Figure 2.

**Figure S.6:**
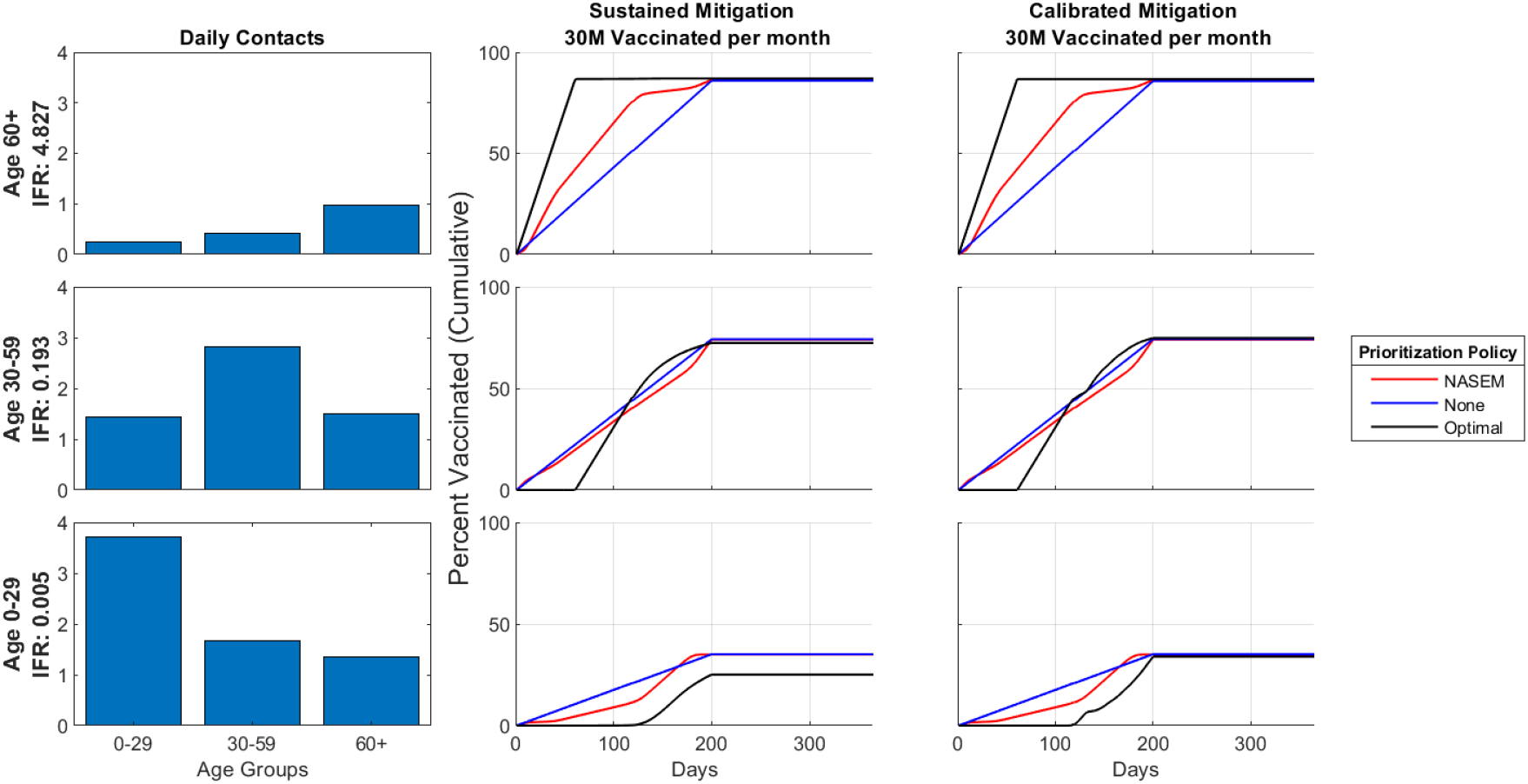
IFR, Contact Rates and Vaccination Policies by Age Group and Mitigation Scenario with Sustained Mitigation set to *ℛ*_0_ = 1.3. Analogous to Figure 1.

**Figure S.7:**
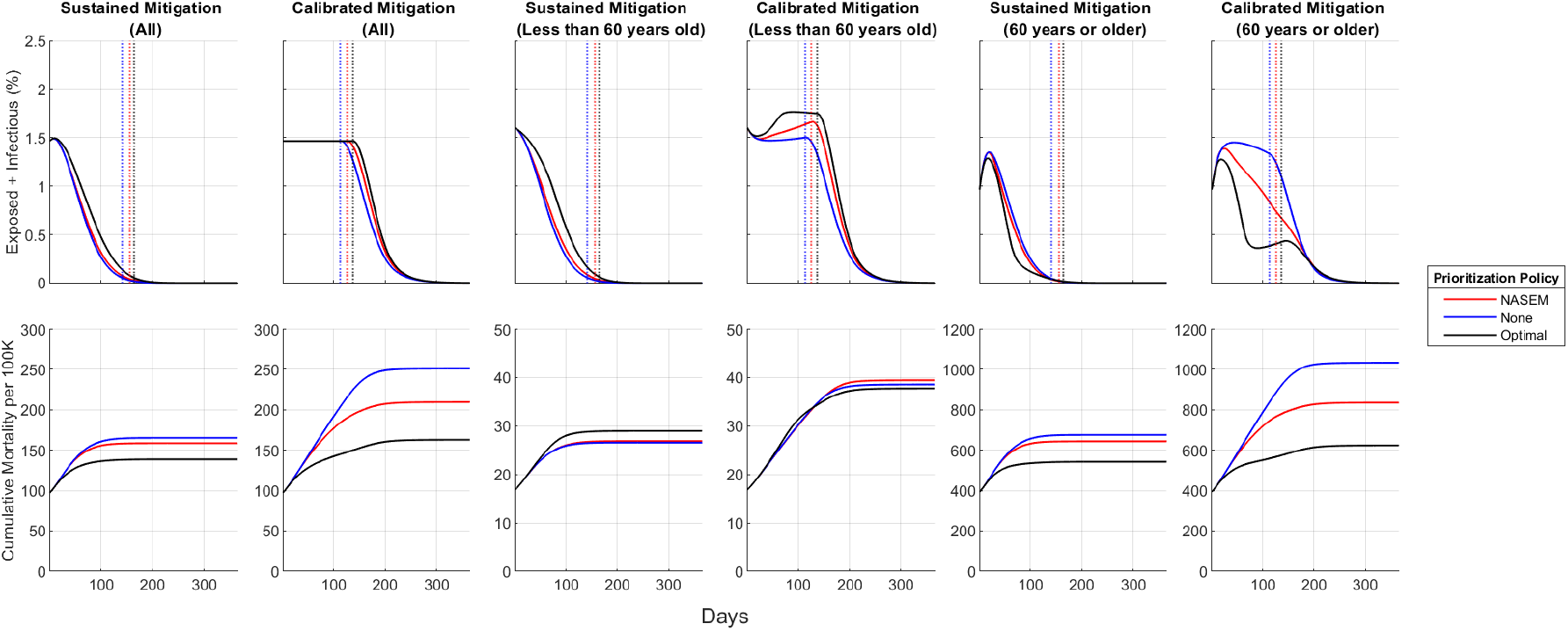
Active Cases and Cumulative Mortality over Time with Sustained Mitigation set to *ℛ*_0_ = 1.3. Analogous to Figure 2.

**Figure S.8:**
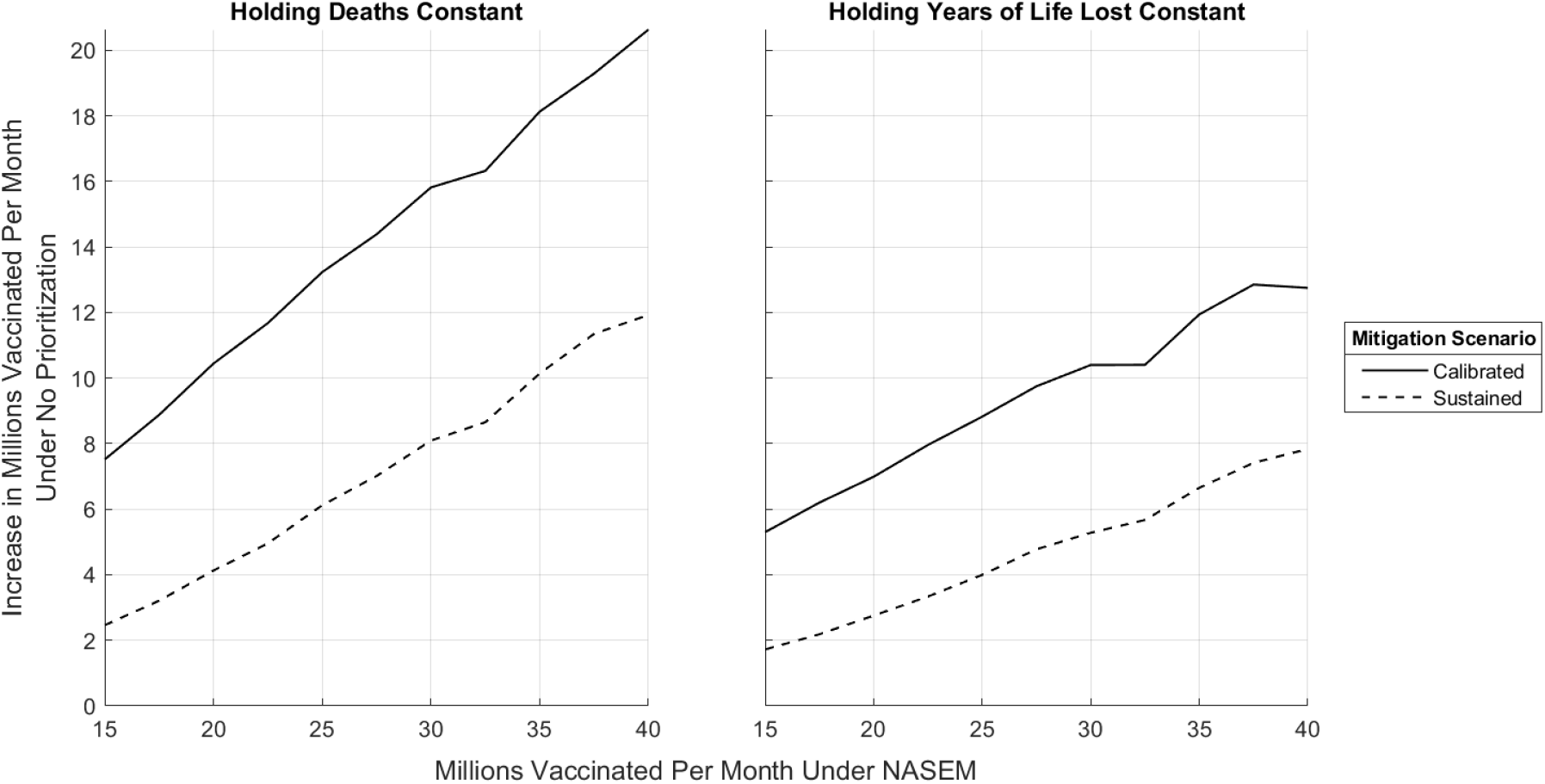
The Required Increase in Vaccinations per month with Sustained Mitigation set to *ℛ*_0_ = 1.3. Analogous to Figure 4.

**Figure S.9:**
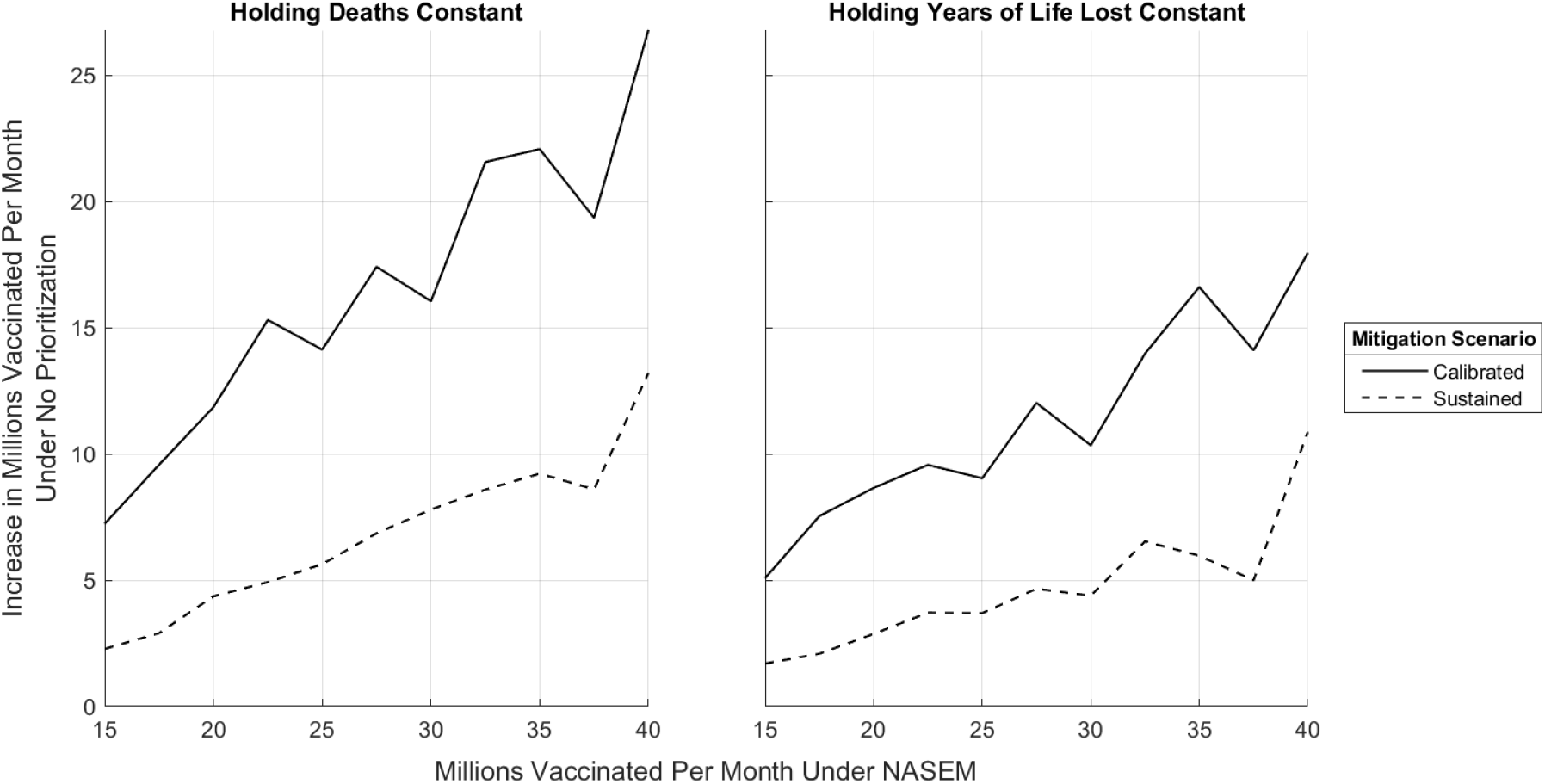
The Required Increase in Vaccinations per month with No Vaccine Hesitancy. Analogous to Figure 4.

**Figure S.10:**
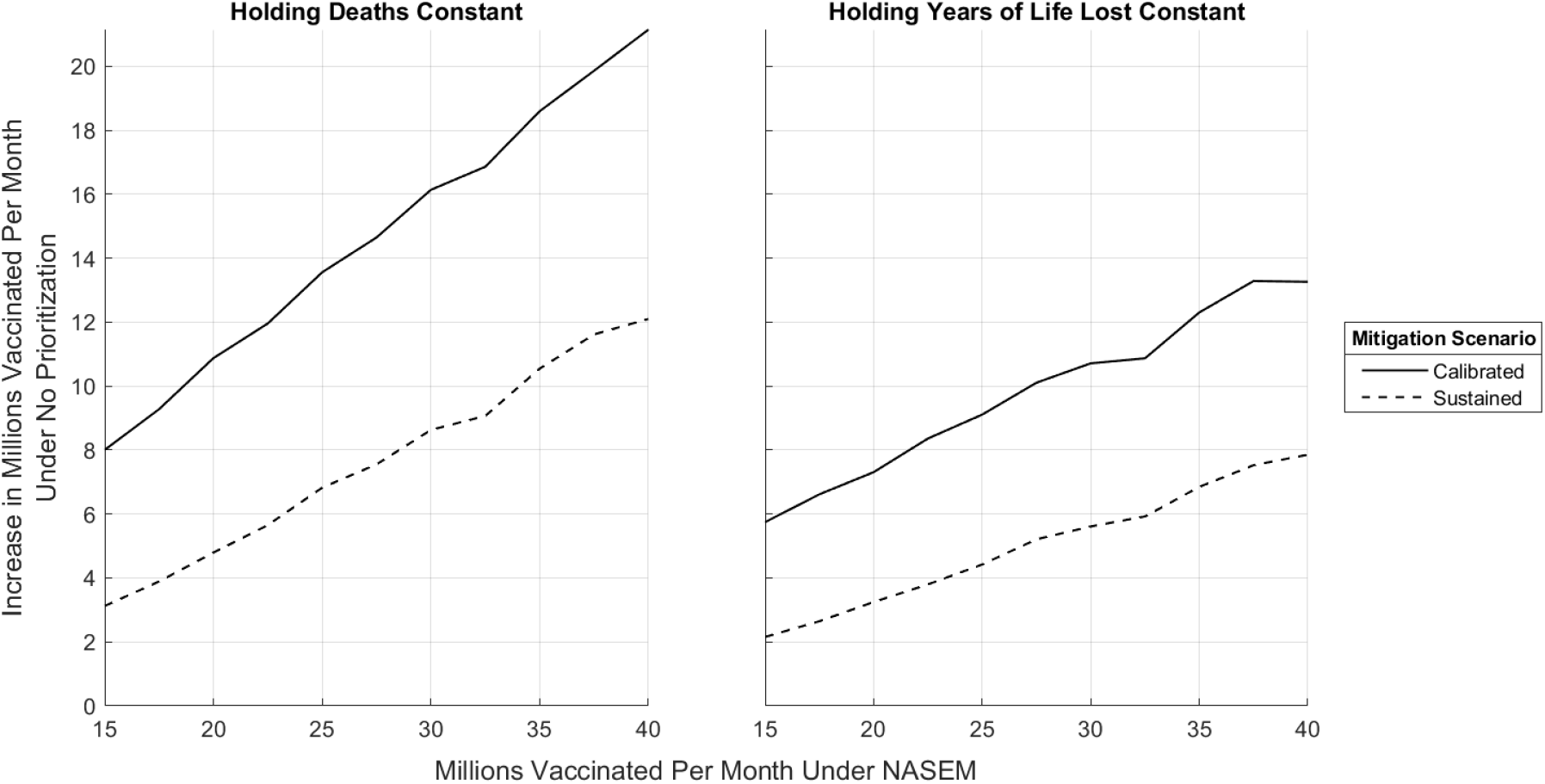
The Required Increase in Vaccinations per month with Vaccine Efficacy set to 80%. Analogous to Figure 4.

**Figure S.11:**
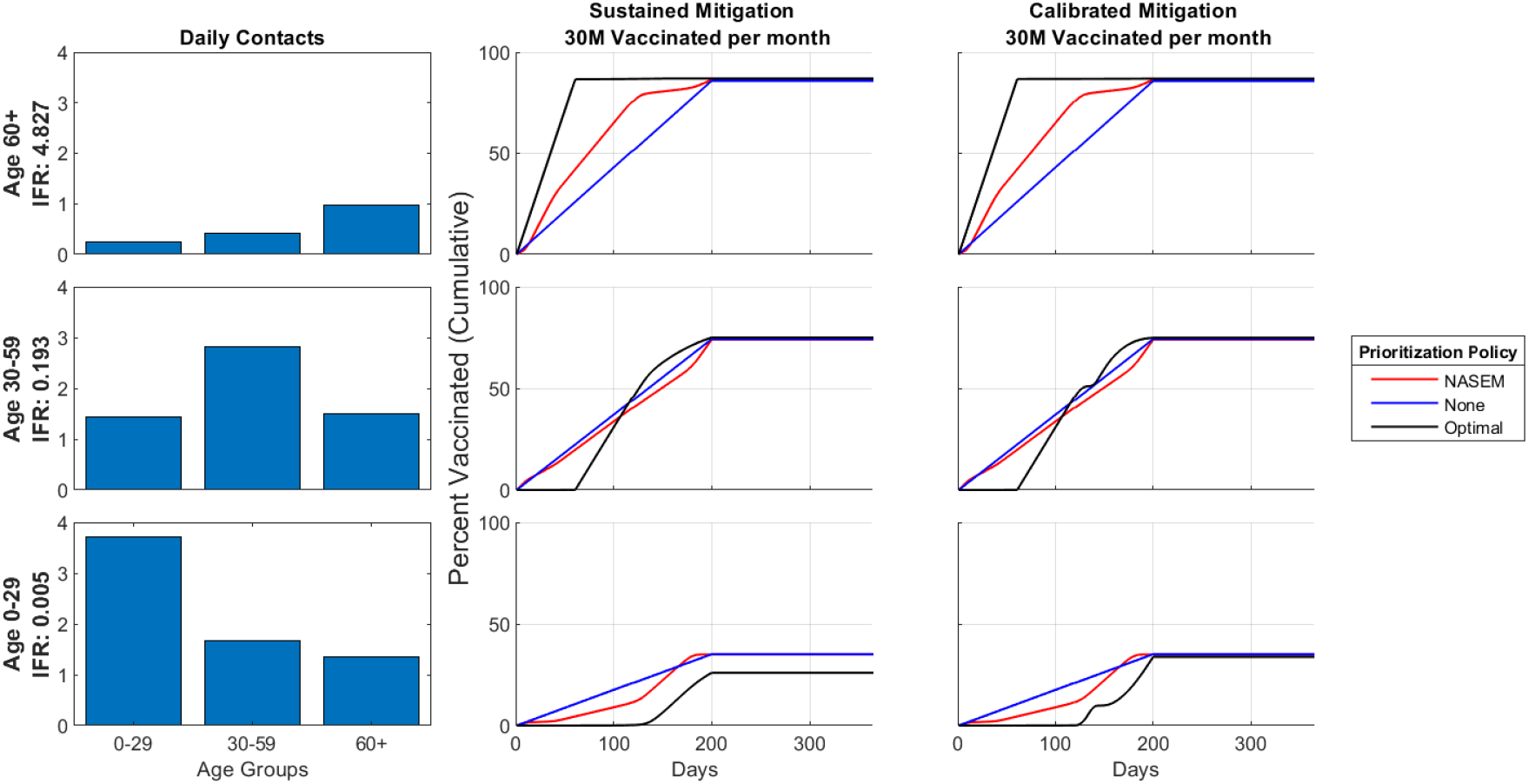
IFR, Contact Rates and Vaccination Policies by Age Group and Mitigation Scenario with Vaccine Efficacy at 80%. Analogous to Figure 1.

**Table S.5:** Age Distribution of Cumulative True Infections. CDC estimates [48] are assumed to include infections occurring through December 31, 2020 whereas our estimates only include infections occurring through December 14, 2020. CDC reported age groups do not align with our group structure.

|  | CDC Estimates |  |  | Our Estimate |
| --- | --- | --- | --- | --- |
|  | Cumul. as of 12/31/20 |  |  | Cumul. as of 12/14/20 |
| **Age** |  |  |  |  |
| 0-17 | 17,552,452 (21.2%) | 0-19 |  | 18,842,982 (28.3%) |
| 18-49 | 41,940,215 (50.5%) | 20-49 |  | 33,133,047 (49.8%) |
| 50+ | 23,486,817 (28.3%) | 50+ |  | 14,499,785 (21.8%) |
| All ages | 83,111,629 | All ages |  | 66,475,815 |

